# Association between intraoperative alactic base excess and postoperative acute kidney injury after liver transplantation: a retrospective cohort study

**DOI:** 10.64898/2026.09.02.26362107

**Authors:** Diego Escarraman Martinez, Salvador Sanchez Diaz, Karen Itzel Pedraza Yañez, Carla Adelina Escorza Molina, Manuel Alberto Guerrero Alberto, Lorena Noriega Salas, Gerardo Alberto Solis Perez, Jorge M. Antolinez-Motta, Alfonso de Jesus Flores Rodriguez

## Abstract

**Background:** Acute kidney injury (AKI) is a common complication after liver transplantation (LT). Alactic base excess (ABE) reflects the metabolic component of acid–base disturbances not attributable to lactate and has been associated with kidney dysfunction in other clinical settings. We evaluated the association between intraoperative ABE and postoperative AKI after LT.

**Methods:** We conducted a retrospective cohort study of adult patients undergoing LT. ABE was calculated from standard base excess and lactate obtained from the last intraoperative arterial blood gas analysis. The primary outcome was creatinine-based AKI occurring within the first 7 postoperative days according to Kidney Disease: Improving Global Outcomes criteria. The association between ABE and AKI was estimated using modified Poisson regression with robust variance, adjusted for age, sex, body mass index, MELD score, cold ischemia time, and anhepatic phase duration. A post hoc exploratory analysis evaluated severe AKI, defined as KDIGO stages 2–3.

**Results:** Seventy patients were included, of whom 32 (45.7%) developed AKI. ABE values were lower among patients who developed AKI than among those who did not (median, −1.15 vs 0.75 mmol/L; Hodges–Lehmann location shift, −1.60 mmol/L; 95%CI, −2.80 to −0.20). After multivariable adjustment, ABE was not independently associated with AKI (aRR, 0.91; 95%CI, 0.82–1.01; p=0.07). Severe AKI occurred in 14 patients (20.0%), and ABE was not independently associated with severe AKI (aRR, 0.98; 95%CI, 0.78–1.23; p=0.87). Sensitivity analyses yielded consistent findings.

**Conclusions:** A single intraoperative ABE measurement was not independently associated with postoperative AKI or severe AKI after LT. Serial measurements across different phases of transplantation may better characterize the potential relationship between ABE dynamics and postoperative kidney injury.

## Introduction

Liver transplantation (LT) represents the only curative treatment option for patients with advanced liver disease [1]. In 2024, 42,497 LT were performed worldwide, based on data collected from 71 countries [2]. In Mexico, during the same year, the National Transplant Center (CENATRA) reported 275 LT [1]. Acute kidney injury (AKI) is one of the major postoperative complications following LT. A systematic review published in 2025, including 30 studies and 13,653 patients, estimated an overall AKI incidence of 46%, with individual study rates ranging from 24% to 84%; approximately 10% of patients required renal replacement therapy [4]. Despite its high incidence and clinical impact, prediction of AKI after LT remains challenging. Several predictive models have been developed; however, their clinical applicability remains limited because of a high risk of bias, limited external validation, and substantial heterogeneity in the variables and definitions used [5].

Alactic base excess (ABE) has been proposed as a marker associated with the risk of developing AKI in patients with sepsis [6,7]. ABE is intended to isolate the metabolic component not attributable to lactate, thereby identifying disturbances related to the accumulation of fixed acids, including those associated with impaired kidney function [8,9]. In addition, ABE may provide complementary information for interpreting acid-base disturbances and could potentially contribute to the individualization of resuscitation strategies [8]. This concept can be explained using Stewart’s physicochemical model: an increase in lactate reduces the apparent strong ion difference (SIDa), thereby promoting metabolic acidosis. When the renal capacity to compensate for this acid load is impaired, fixed acids or unmeasured anions other than lactate may accumulate, a phenomenon reflected by more negative ABE values [7,8]. ABE may help distinguish the lactate-related component from the non-lactate metabolic component of acidosis and provide additional information regarding the contribution of renal function to the acid-base disorder.

Despite its biological plausibility and the associations reported between ABE and AKI in other clinical settings, such as sepsis, its role during LT has not been investigated. LT is characterized by marked hemodynamic and metabolic disturbances, including substantial fluctuations in blood pressure, acidosis, ischemia-reperfusion injury, dynamic changes in lactate concentrations and acid-base balance, and considerable variations in fluid requirements [10,11]. All of these factors may contribute to the development of AKI and may also influence the interpretation of intraoperative ABE. Therefore, we hypothesized that intraoperative ABE would be associated with the development of AKI during the first 7 postoperative days in patients undergoing LT.

## Materials and Methods

### Study Design and Ethical Considerations

A retrospective cohort study was conducted at Hospital de Especialidades “La Raza” in Mexico City.The source population comprised patients who underwent LT at our institution during a predefined 3-year inclusion period. The study itself, including protocol development and approval, data collection, and statistical analysis, was conducted over a 13-month period. The study was approved by the Comité Local de Investigación en Salud (CLIS-R2026-3501-069). The analysis protocol was registered in AsPredicted before the statistical analysis was performed (267497). To protect participant identity and preserve the confidentiality of clinical information, each included patient was assigned a unique alphanumeric identifier. The dataset used for analysis was deidentified by removing direct personal identifiers before deposition in a public repository. The anonymized dataset will be made available through Harvard Dataverse with a persistent Digital Object Identifier (DOI). The study was conducted in accordance with the principles of the Declaration of Helsinki and the Declaration of Istanbul on Organ Trafficking and Transplant Tourism. The requirement for informed consent was waived by the ethics committee because of the retrospective nature of the study. The study was reported in accordance with the Strengthening the Reporting of Observational Studies in Epidemiology (STROBE) statement and its extension for studies conducted using routinely collected health data, the REporting of studies Conducted using Observational Routinely-collected health Data (RECORD) statement [12,13].

### Study Population and Variables

We included male and female patients aged >18 years who underwent LT during the inclusion period and had complete follow-up during the first 7 postoperative days, including serum creatinine measurements and urine output monitoring. Patients with incomplete clinical information, those undergoing retransplantation, and those who died within the first 7 postoperative days were excluded. Collected variables included age (years), sex, body mass index (BMI, kg/m²), cold ischemia time (hours), duration of the anhepatic phase (hours), etiology of liver disease, use and type of diuretics, use and type of vasopressors, Model for End-Stage Liver Disease (MELD) score, surgical technique (Piggyback or portocaval), intraoperative blood loss (L), and fluid balance (L). The exposure was ABE, calculated according to the formula proposed by Gattinoni et al. [8]: ABE = standard base excess (mmol/L) + lactate (mmol/L). Both components were obtained from the last intraoperative arterial blood gas analysis documented in the anesthesia record. Intraoperative blood loss and fluid balance were obtained from the same records. The remaining variables were collected from medical records documented during the ICU stay. Data were extracted directly from the clinical records by a single anesthesiology resident.

### Outcomes

The primary outcome was the development of AKI during the first 7 postoperative days, defined according to the serum creatinine component of the Kidney Disease: Improving Global Outcomes (KDIGO) criteria [14]. Baseline serum creatinine was defined as the day 0 value. For severity classification, creatinine-based KDIGO stages were assigned according to the maximum serum creatinine change observed during the first 7 postoperative days. Stage 1 was defined as an increase in serum creatinine to 1.5–1.9 times baseline or an increase of ≥0.3 mg/dL within 48 hours; stage 2 as an increase to 2.0–2.9 times baseline; and stage 3 as an increase to ≥3.0 times baseline or a serum creatinine concentration ≥4.0 mg/dL. Patients not meeting any of these criteria were classified as KDIGO stage 0. Urine output was recorded during the postoperative period but was not used for KDIGO severity staging because it was available as daily summarized values and did not allow reliable assessment of the specific duration thresholds required by KDIGO. The association between intraoperative ABE and the development of AKI constituted the primary analysis. As an exploratory secondary outcome, time to ICU discharge during the first 7 postoperative days was evaluated.

### Sample Size

Given the retrospective design of the study, no formal a priori sample size calculation was performed. To characterize the accessible population, the volume of liver transplants performed at the institution during the preceding 3 years was reviewed. The analytical cohort, however, included all patients who underwent LT during the predefined study period and met the eligibility criteria. Therefore, a census sampling approach was used for the eligible population during that period. After application of the exclusion criteria, the final cohort comprised 70 patients. The precision of the association estimates was assessed using 95% confidence intervals (95%IC).

### Statistical Analysis

The distribution of continuous variables was assessed using the Shapiro–Wilk test, with a p-value >0.05 considered compatible with a normal distribution. Quantitative variables were summarized as median and interquartile range (IQR), whereas categorical variables were presented as absolute frequencies and percentages. To describe differences between groups (AKIi vs no AKI), location shifts for continuous variables were estimated using the Hodges–Lehmann estimator. For categorical variables, differences in proportions were calculated. Both estimates were reported with their corresponding 95% confidence intervals (95%CI).

Before multivariable analysis, a directed acyclic graph (DAG) was developed using clinical knowledge and biological plausibility to explicitly represent the hypothesized causal relationships between ABE and postoperative AKI. Potential confounding was addressed using a prespecified adjustment set for the primary multivariable analysis, informed by the causal structure represented in the DAG, clinical knowledge, biological plausibility, and considerations regarding model complexity. Covariate selection was not based on statistical significance testing [15]. The DAG was constructed and analyzed using the *dagitty* package in R [16]. The primary adjustment set included age, sex, BMI, MELD score, cold ischemia time, and anhepatic phase duration. Given the binary nature of the outcome, the association between ABE and AKI was estimated using modified Poisson regression with robust variance [17]. This approach was selected to obtain direct estimates of the adjusted risk ratio (aRR), because odds ratios derived from logistic regression may differ substantially from risk ratios and overstate the magnitude of an association when the outcome is not uncommon [17,18].

Before final model adjustment, collinearity among covariates was assessed using the variance inflation factor (VIF), with values >5 considered indicative of relevant collinearity. Because ABE was analyzed as a continuous variable, the functional form of its association with the outcome was explored using restricted cubic splines with three knots to assess potential deviations from linearity [19]. The linear specification was compared with the spline model using the Akaike information criterion (AIC) and a chi-square test based on the difference in deviance between nested models. In the absence of evidence of a nonlinear association, the linear specification was retained for the primary analysis. Results were reported as aRR with their corresponding 95%CI.

Two sensitivity analyses were performed. First, ABE was dichotomized at −2.5 mmol/L and its association with AKI was re-estimated using the same modified Poisson model with robust variance and the same adjustment set. Second, potential effect modification by anhepatic phase duration was explored by including an ABE × anhepatic phase multiplicative interaction term in the regression model. For the exploratory analysis of time to ICU discharge, Kaplan–Meier curves were constructed and stratified according to ABE groups (≤−2.5 mmol/L vs >−2.5 mmol/L). Patients who remained in the ICU beyond postoperative day 7 were censored at that time point, and differences between curves were assessed using the log-rank test.

As a post hoc exploratory analysis, AKI severity was examined according to creatinine-based KDIGO stages 0, 1, 2, and 3. The distribution of ABE across KDIGO stages was summarized using medians and IQRs and compared using the Kruskal–Wallis test. Severe AKI was subsequently defined as KDIGO stages 2–3, whereas patients with KDIGO stages 0–1 constituted the reference group. The association between continuous ABE and severe AKI was estimated using modified Poisson regression with robust variance. Both unadjusted and adjusted estimates were obtained, with the adjusted model using the same a priori covariate set as the primary analysis. As an additional exploratory analysis, ABE dichotomized at −2.5 mmol/L was evaluated in relation to severe AKI using the same adjusted model. Given the post hoc nature of these analyses, results were considered exploratory and hypothesis-generating.

All statistical tests were two-sided, and a p-value <0.05 was considered statistically significant for the primary and prespecified analyses. No adjustment for multiple comparisons was applied to the post hoc exploratory analyses; therefore, their p-values and 95%CI were interpreted descriptively rather than as confirmatory evidence. Robust 95%CI for modified Poisson models were calculated using heteroskedasticity - consistent (HC0) standard errors. Statistical analyses were performed using R (R Foundation for Statistical Computing, Vienna, Austria) through RStudio (Posit Software, Boston, MA, USA).

### Bias Control

To reduce potential sources of bias, uniform eligibility criteria were applied to patients undergoing LT during the study period. Data collection was performed by a single investigator using predefined sources within the clinical records. Intraoperative variables were obtained from anesthesia records, whereas postoperative variables were collected from clinical and nursing records during the ICU stay. To reduce the risk of exposure misclassification, ABE was calculated consistently in all patients using the validated formula, with standard base excess and lactate obtained from the same final intraoperative arterial blood gas analysis. The primary outcome was defined using standardized KDIGO criteria, with a uniform definition of baseline creatinine and urine output derived from nursing records. Potential confounding was addressed through the a priori construction of a DAG based on clinical knowledge and biological plausibility, from which a minimally sufficient adjustment set was identified. Covariate selection was not based on statistical significance testing.

## Results

A total of 95 clinical records were reviewed. Of these, 16 patients were excluded because of incomplete clinical information, 7 because of death within the first 7 postoperative days, and 2 because they underwent retransplantation, resulting in a final cohort of 70 patients for analysis. The median age was 49.5 years (IQR, 40–56), and 49% of patients were female. NAFLD was the most common etiology of liver disease (63%), and the median MELD score was 18 (IQR, 15–21). Creatinine-based AKI occurred in 32 patients (45.7%). Median ABE was −0.50 mmol/L (IQR, −2.70 to 2.20), and median ICU length of stay was 7 days (IQR, 5–9). Additional baseline and perioperative characteristics are summarized in Table 1. Patient comparison (No AKI vs AKI), ABE values were lower among patients who developed AKI versus no AKI (−1.15 mmol/L [IQR, −3.02 to 0.85] vs 0.75 mmol/L [IQR, −1.53 to 2.85]), corresponding to a Hodges–Lehmann location shift estimate of −1.60 mmol/L (95% CI, −2.80 to −0.20). MELD scores were also higher among patients with AKI (20 [IQR, 17–22] vs 17 [IQR, 13.25–19.75]), with a Hodges–Lehmann location shift estimate of 3 points (95%CI, 0– 5). No clear between-group differences were observed for the remaining variables. Full comparisons are presented in Table 2.

**Table 1.** General description of the study variables. NAFLD: Non-Alcoholic Fatty Liver Disease; MELD: Model for End-Stage Liver Disease; ICU: intensive care unit. * median (interquartile range); ** frequency (percentage)

| Characteristic | N = 70 |
| --- | --- |
| Age, years* | 49.50 (40.00-56.00) |
| Sex, Female** | 34 (49) |
| Body mass index, kg/m<sup>2</sup>* | 25.45 (22.80-28.00) |
| Cold ischemia time, h* | 7.29 (6.66-8.00) |
| Anhepatic phase duration, h* | 1.77 (1.25-3.11) |
| Fluid balance, L* | 0.21 (-1.05-1.21) |
| MELD score* | 18.00 (15.00-21.00) |
| Intraoperative blood loss, L* | 3.60 (2.50-5.50) |
| Etiology of liver disease** |  |
| NAFLD | 44 (63) |
| Alcohol-related | 7 (10) |
| Hepatitis | 19 (27) |
| Diuretic use** | 41 (59) |
| Diuretic type** |  |
| Furosemide | 19 (27) |
| Mannitol | 7 (10) |
| Furosemide + Mannitol | 15 (21) |
| Vasopressor use** | 67 (96) |
| Vasopressor type** |  |
| Norepinephrine | 38 (54) |
| Norepinephrine + Vasopressin | 29 (41) |
| Surgical technique** |  |
| Piggyback | 52 (74) |
| Portocaval | 18 (26) |
| Alactic base excess, mmol/L* | -0.50 (-2.70-2.20) |
| Acute kidney injury** | 32 (45.7) |
| ICU length of stay, days* | 7.00 (5.00-9.00) |

**Table 2.** Comparison between groups according to acute kidney injury (AKI) status: NAFLD: non-alcoholic fatty liver disease; MELD: Model for End-Stage Liver Disease; ICU: intensive care unit; 95%IC: 95% confidence interval. * Hodges–Lehmann estimator. ** Difference in proportions.

| Characteristic | No AKI (n=38) | AKI (n=32) | Effect estimate (95%CI) |
| --- | --- | --- | --- |
| Age, years* | 50.5 (40-58.75) | 47.5 (41-53.25) | -2.00 (-8.00 to 3.00 ) |
| Sex, Female** | 20 (52.6%) | 14 (43.8%) | -8.88 (-32.3 to 14.5) |
| Body mass index, kg/m<sup>2</sup>* | 25.25 (22.58-27.2) | 25.85 (23.67-28.42) | 1.10 (-0.80 to 3.00) |
| Cold ischemia time, h* | 7.07 (6.45-7.89) | 7.38 (6.94-8) | 0.198 (-0.350 to 0.670) |
| Anhepatic phase duration, h* | 1.64 (1.23-2.66) | 2.16 (1.29-3.25) | 0.211 (-0.240 to 0.730) |
| Fluid balance, L* | 0.37 (-0.78-1.79) | 0.03 (-1.43-0.59) | -0.777 (-1.87 to 0.190) |
| MELD score* | 17 (13.25-19.75) | 20 (17-22) | 3.0 (0.000057 to 5.00) |
| Intraoperative blood loss, L* | 3.26 (2.5-5) | 4.52 (2.45-6.38) | 1.00 (-0.280 to 2.30) |
| Etiology of liver disease** |  |  |  |
| NAFLD | 24 (63.2%) | 20 (62.5%) | -0.658 (-23.4 to 22.1) |
| Alcohol-related | 2 (5.3%) | 5 (15.6%) | 10.4 (-4.08 to 24.8) |
| Hepatitis | 12 (31.6%) | 7 (21.9%) | -9.70 (-30.3 to 10.9) |
| Diuretic use** | 20 (52.6%) | 21 (65.6%) | 13.0 (-9.87 to 35.9) |
| Diuretic type** |  |  |  |
| Furosemide | 9 (23.7%) | 10 (31.2%) | 7.57 (-13.4 to 28.6) |
| Mannitol | 2 (5.3%) | 5 (15.6%) | 10.4 -4.08 to 24.8) |
| Furosemide + Mannitol | 9 (23.7%) | 6 (18.8%) | -4.93 (-24.1 to 14.2) |
| Vasopressor use** | 36 (94.7%) | 31 (96.9%) | 2.14 (-7.18 to 11.5) |
| Vasopressor type** |  |  |  |
| Norepinephrine | 24 (63.2%) | 14 (43.8%) | -19.4 (-42.4 to 3.63) |
| Norepinephrine + Vasopressin | 12 (31.6%) | 17 (53.1%) | 21.5 (-1.20 to 44.3) |
| Surgical technique** |  |  |  |
| Piggyback | 29 (76.3%) | 23 (71.9%) | -4.44 (-25.1 to 16.2) |
| Portocaval | 9 (23.7%) | 9 (28.1%) | 4.44 (-16.2 to 25.1) |
| Alactic base excess, mmol/L* | 0.75 (-1.53-2.85) | -1.15 (-3.02-0.85) | -1.60 (-2.80 to -0.20) |
| ICU length of stay, days* | 6.5 (5-9) | 8 (5-10) | 1.00 (-1.00 to 2.00) |

The prespecified adjustment set comprised age, sex, BMI, MELD score, cold ischemia time, and anhepatic phase duration, with the hypothesized causal relationships among relevant variables represented in Figure 1. VIF values were low for age (1.25), sex (1.47), BMI (1.20), MELD score (1.23), cold ischemia time (1.12), anhepatic phase duration (1.20), and ABE (1.25), indicating no relevant collinearity among the variables included in the model. A chi-square test comparing the linear and restricted cubic spline models showed no evidence of nonlinearity in the association between ABE and AKI (p=0.79). The linear model also yielded a lower AIC than the restricted cubic spline model (125.44 vs 127.37); therefore, the linear specification was retained for the primary analysis. In the primary modified Poisson regression with robust variance, ABE was not independently associated with postoperative AKI (aRR, 0.91; 95% CI, 0.82– 1.01; p=0.07). No clear evidence of association with AKI was observed for the remaining covariates. Full results are presented in Table 3. The absence of clear evidence of an association between ABE and AKI was consistent across sensitivity analyses. When ABE was dichotomized at −2.5 mmol/L, ABE ≤−2.5 mmol/L was not independently associated with AKI (aRR, 1.42; 95% CI, 0.85–2.38; p=0.19). Likewise, there was no evidence of effect modification by anhepatic phase duration (ABE × anhepatic phase duration interaction aRR, 1.00; 95% CI, 0.91–1.10; p=0.98) (Supplemental Table 1-2). In the exploratory analysis of time to ICU discharge. Kaplan–Meier curves showed a similar pattern between ABE groups during the first 7 postoperative days, with no evidence of a difference between the curves (log-rank p=0.62) (Figure 2).

**Figure 1.**
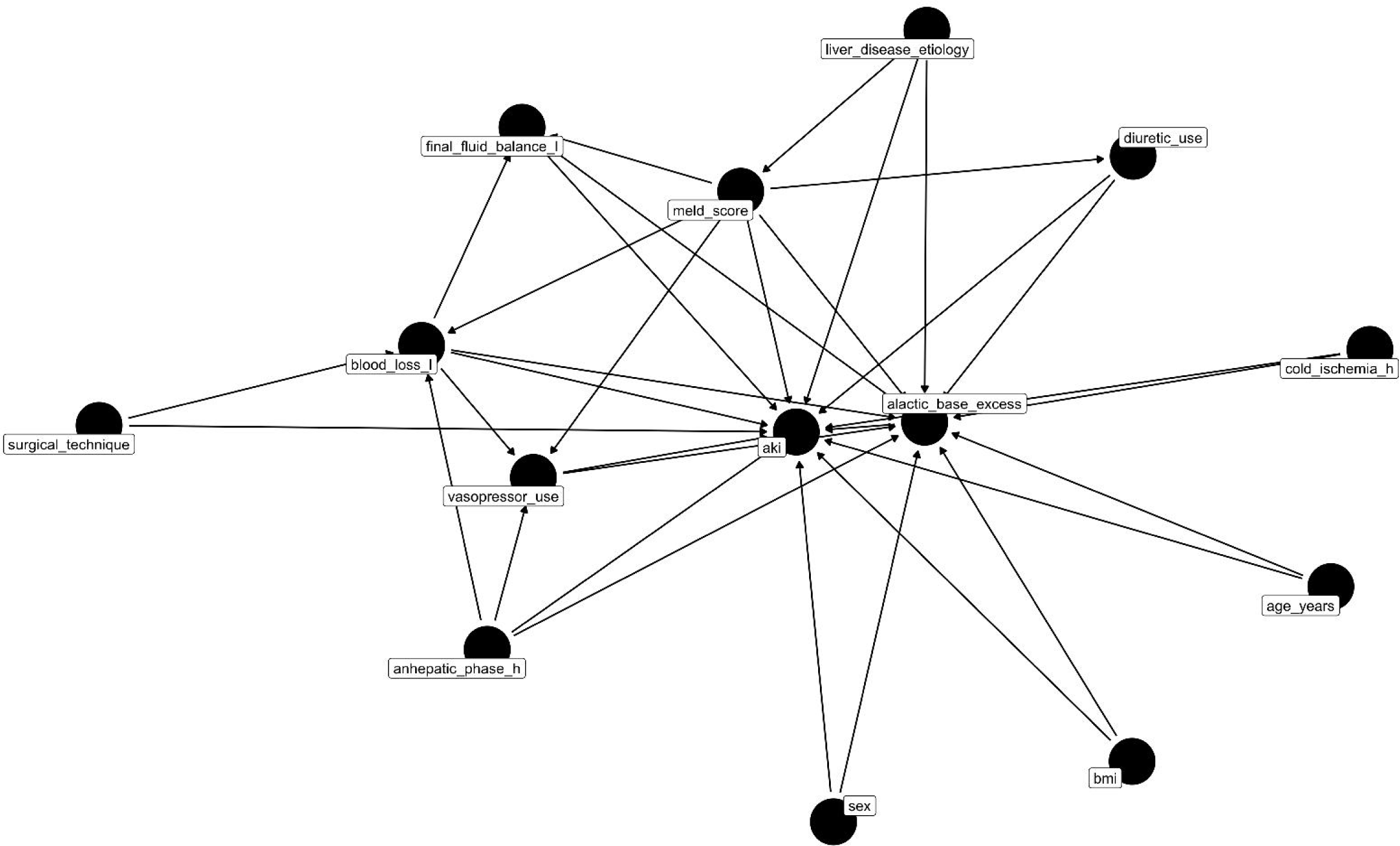
Directed acyclic graph (DAG) representing the hypothesized causal relationships between intraoperative alactic base excess (ABE) and postoperative acute kidney injury (AKI) in patients undergoing liver transplantation. The DAG was constructed based on biological plausibility and prior clinical knowledge to identify the minimally sufficient adjustment set for the multivariable analysis. Arrows represent the assumed causal relationships among the covariates included in the model. MELD: Model for End-Stage Liver Disease; AKI: acute kidney injury; BMI: body mass index.

**Figure 2.**
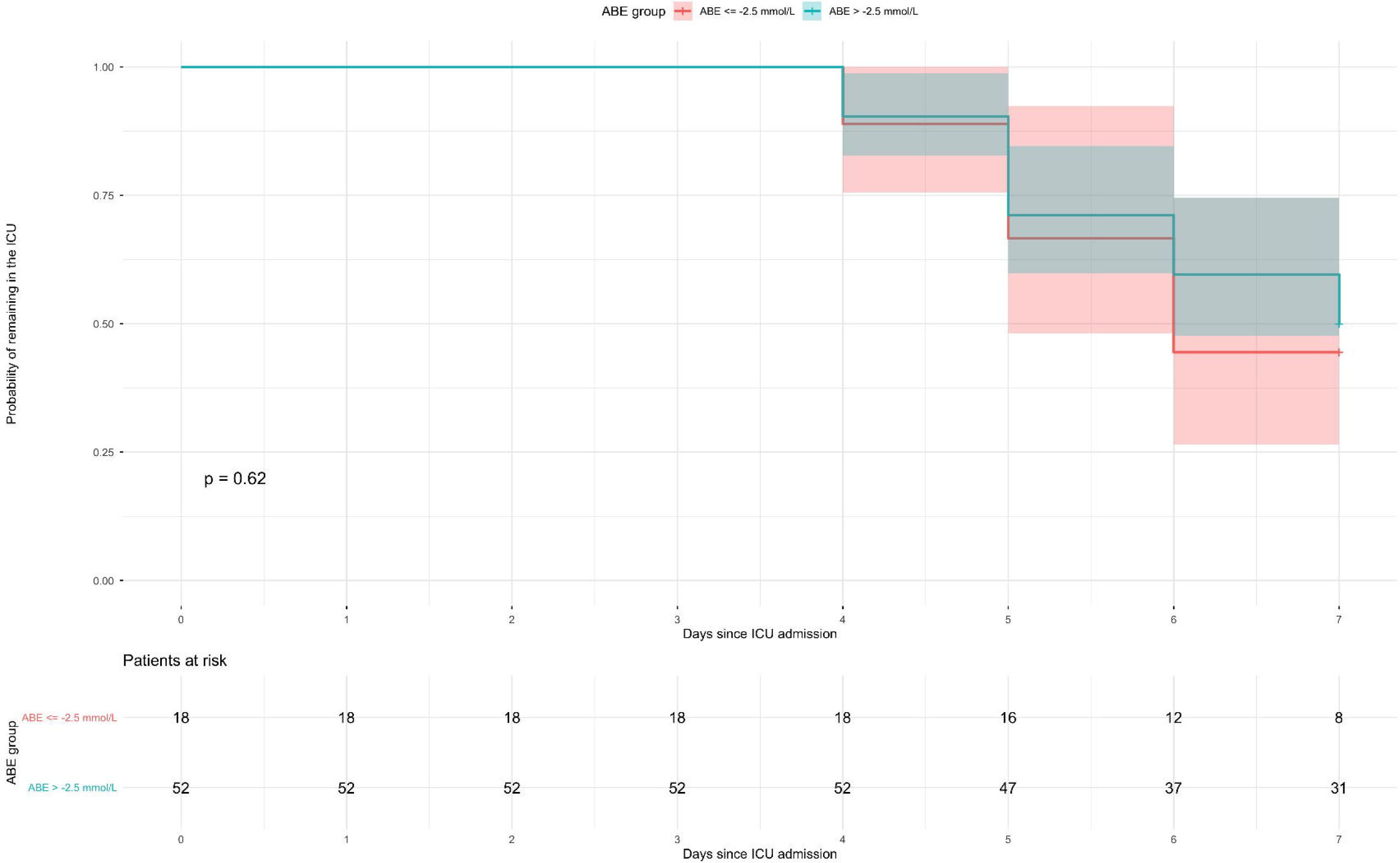
Kaplan–Meier curves for time to intensive care unit (UCI) discharge, defined as an UCI stay of ≤7 days, stratified according to alactic base excess (ABE) groups. Differences between curves were compared using the log-rank test.

**Table 3.**
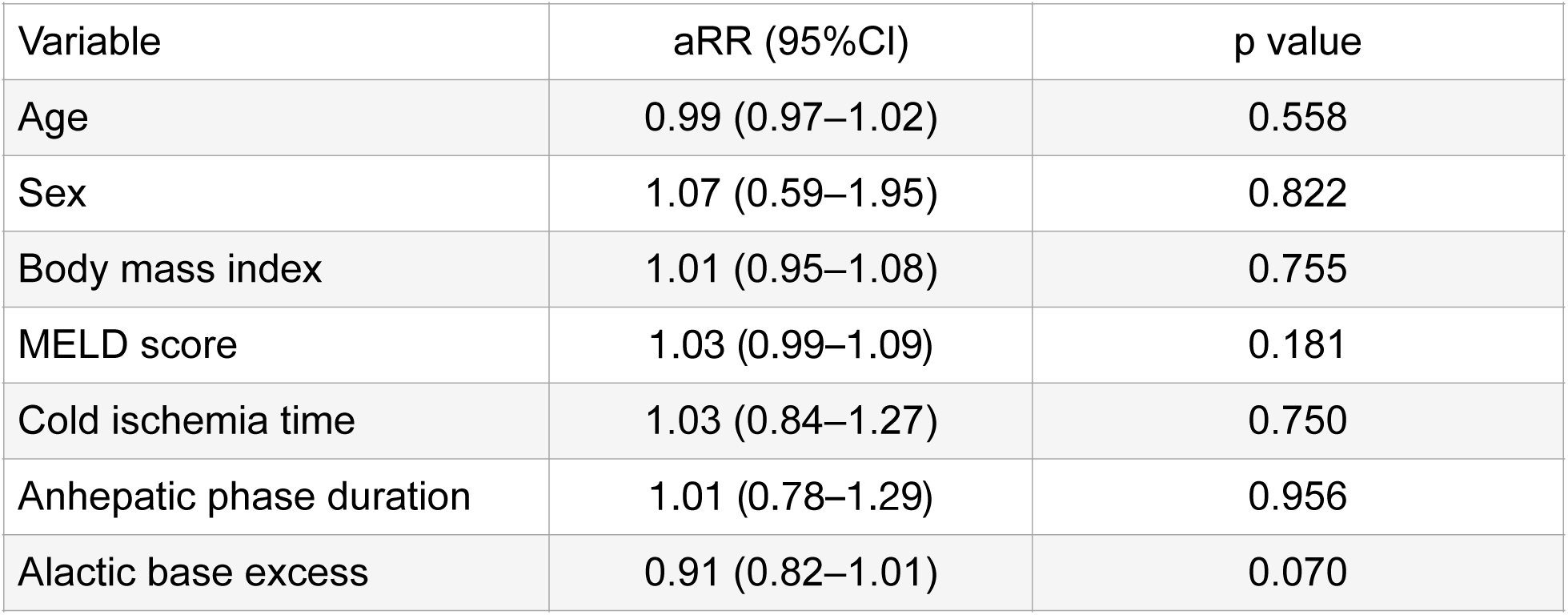
Modified Poisson regression with robust variance. MELD: Model for End-Stage Liver Disease; aRR: adjusted relative risk; 95%CI: 95% confidence interval.

| Variable | aRR (95%CI) | p value |
| --- | --- | --- |
| Age | 0.99 (0.97–1.02) | 0.558 |
| Sex | 1.07 (0.59–1.95) | 0.822 |
| Body mass index | 1.01 (0.95–1.08) | 0.755 |
| MELD score | 1.03 (0.99–1.09) | 0.181 |
| Cold ischemia time | 1.03 (0.84–1.27) | 0.750 |
| Anhepatic phase duration | 1.01 (0.78–1.29) | 0.956 |
| Alactic base excess | 0.91 (0.82–1.01) | 0.070 |

In the post hoc exploratory analysis of AKI severity, 38 patients (54.3%) were classified as KDIGO stage 0, 18 (25.7%) as stage 1, 8 (11.4%) as stage 2, and 6 (8.6%) as stage 3. ABE distributions differed across KDIGO stages (Kruskal–Wallis p=0.028); however, no monotonic pattern was observed with increasing AKI severity. Severe AKI, defined as KDIGO stages 2–3, occurred in 14 patients (20.0%). In the adjusted modified Poisson model, ABE was not independently associated with severe AKI (aRR, 0.98; 95%CI, 0.78–1.23; p=0.87). (Supplemental Table 3-4)

## Discussion

In this study, lower intraoperative ABE values were observed among patients who developed AKI; however, this association was attenuated after adjustment for prespecified covariates, and ABE was not independently associated with postoperative AKI. Although the adjusted estimate was compatible with a modest reduction in AKI risk with increasing ABE, the 95%CI included the null value. These findings suggest that a single intraoperative ABE measurement may not, by itself, adequately capture the complexity of the mechanisms involved in the development of AKI after LT.

The use of a DAG allowed this association to be evaluated while explicitly accounting for potential confounding factors and avoiding unnecessary adjustment for variables that might lie on the causal pathway. Our findings are consistent with the evidence describing AKI after LT as a multifactorial phenomenon. Berkowitz et al. [20] identified several preoperative and intraoperative factors associated with AKI, including fluid, electrolyte, and acid-base disturbances, anemia, lower albumin concentrations, and alterations in potassium and lactate levels during reperfusion. These findings suggest that metabolic abnormalities may contribute to the development of AKI, but likely as part of a dynamic interaction with the clinical, hemodynamic, and metabolic conditions surrounding transplantation. Similarly, Tedesco et al. [21], in a systematic review of the available evidence, highlighted the contribution of multiple potentially modifiable perioperative factors to the development of AKI after LT, including hemodynamic instability, metabolic disturbances during reperfusion, transfusion requirements, and several components of intraoperative anesthetic management. Taken together, these findings reinforce the multifactorial nature of AKI after LT and make it unlikely that a single metabolic marker can independently capture perioperative renal risk.ABE has a close pathophysiological relationship with renal function and acid load handling. The kidneys contribute to acid-base homeostasis through the excretion of nonvolatile acids and regeneration of bicarbonate; when this capacity is impaired, retention of fixed acids and unmeasured anions may contribute to more negative ABE values. Gattinoni et al. [8] observed in patients with sepsis that hyperlactatemia was accompanied by acidemia mainly when renal dysfunction coexisted, a condition that could be identified by a negative ABE.

In this sense, ABE provides a conceptual means of separating the contribution of lactate from other metabolic components of the acid load. This interpretation is particularly relevant in the perioperative setting. Using a physicochemical approach in patients undergoing cardiac surgery, Guarnieri et al. [22] demonstrated a substantial contribution of unmeasured anions to postoperative metabolic acidosis, independent of hyperlactatemia. ABE also identified nonlactate metabolic abnormalities that persisted even as pH recovered. These findings support the assessment of metabolic components beyond lactate to more accurately characterize perioperative acid-base disturbances. At the same time, they suggest that a negative ABE should not be regarded as a specific marker of kidney injury, but rather as an integrated expression of metabolic acid burden not explained by lactate. In other clinical settings, lower ABE values have been associated with impaired renal function. Li et al. [23], in patients undergoing coronary artery bypass grafting, found that lower ABE values were independently associated with both worsening renal function and the development of de novo AKI. These findings support the possibility that the metabolic component not explained by lactate may provide additional information on renal risk in selected perioperative settings. Similar findings have been reported in patients with sepsis. Mouli et al. [24] identified baseline ABE as an independent predictor of AKI, with progressively less negative values associated with a lower risk. However, this association appeared to depend on the timing of measurement: although ABE measured at admission retained independent predictive value, measurements obtained at 12 and 24 hours did not. This observation is particularly relevant to the interpretation of our findings, as it suggests that the clinical utility of ABE may depend not only on its magnitude but also on the timing of its assessment.

The consistency of the findings across sensitivity analyses further supports the primary results. Neither dichotomization of ABE (−2.5 mmol/L) nor assessment of its interaction with anhepatic phase duration provided evidence of an independent association with AKI. Evidence from patients with shock provides additional, although indirect, insight. Smuszkiewicz et al. [25] found that markedly negative ABE values at ICU admission were independently associated with higher 28-day mortality. However, because mortality rather than AKI was the outcome of interest, these findings should be interpreted as evidence of the prognostic value of ABE in critically ill patients rather than as evidence of a specific relationship with kidney injury. Taken together, these differences suggest that the clinical meaning of ABE may be context dependent. In sepsis, shock, or cardiac surgery, a negative ABE may reflect a persistent metabolic disturbance within a relatively defined pathophysiological setting. In contrast, LT is characterized by the simultaneous occurrence of rapid changes in lactate metabolism and clearance, ischemia-reperfusion injury, bleeding, transfusion, fluid administration, and hemodynamic instability. Therefore, a single ABE measurement may represent only a snapshot of a highly dynamic metabolic process, potentially limiting its ability to discriminate the subsequent risk of AKI. The post hoc analysis of AKI severity yielded a similar interpretation: although ABE distributions differed across creatinine-based KDIGO stages, no monotonic gradient was observed with increasing severity, and ABE was not independently associated with severe AKI.

This study has several important strengths. The use of a DAG allowed the a priori definition of a minimally sufficient adjustment set based on clinical knowledge and biological plausibility, avoiding covariate selection based on statistical significance. ABE was analyzed as a continuous variable, potential nonlinearity was formally assessed, and modified Poisson regression with robust variance was used to directly estimate risk ratios. AKI was defined using standardized KDIGO criteria, and the robustness of the findings was explored through sensitivity analyses. In addition, several limitations should be acknowledged. First, the relatively small cohort and limited number of AKI events, determined by the eligible population available during the study period, limited the precision of the estimates and the complexity of the multivariable analyses. Although the adjusted estimate for ABE was compatible with a modest inverse association with AKI, 95%CI included the null value; therefore, a small association in either direction cannot be excluded. Second, the retrospective, single-center design limits the generalizability of the findings and leaves the possibility of information bias and residual confounding. Third, AKI and its severity were classified using the serum creatinine component of the KDIGO criteria. Because urine output was available only as daily summarized values, the specific duration thresholds required for KDIGO staging could not be reliably assessed, potentially leading to underestimation or misclassification of AKI severity. The exclusion of patients who died within the first 7 postoperative days may also have introduced selection or survivor bias, particularly if these patients were at higher risk of AKI. In addition, ABE was calculated from a single intraoperative arterial blood gas analysis, precluding assessment of its temporal trajectory and variation across the different phases of LT. Finally, the post hoc analysis of severe AKI was based on only 14 events; therefore, these estimates were imprecise and should be interpreted as exploratory and hypothesis-generating.

## Conclusion

In this retrospective cohort of patients undergoing LT, ABE was not independently associated with postoperative AKI after multivariable adjustment. These findings suggest that a single ABE measurement at the end of the intraoperative period may have limited ability to independently characterize renal risk in a pathophysiologically complex and dynamic setting such as LT. Larger prospective studies incorporating serial ABE measurements across different phases of transplantation are needed to determine whether its temporal trajectory provides greater clinical value for AKI risk stratification.

## Data Availability

The deidentified dataset underlying the findings of this study is available from the corresponding author upon reasonable request and will be deposited in Harvard Dataverse. The persistent DOI and repository link will be added once available.

## Author Contributions (CRediT)

Conceptualization: SSD. Data curation: DEM. Formal analysis: DEM. Investigation: KIPY, GASP. Methodology: MAGA. Project administration: CAEM. Supervision: JMAM. Visualization: LNS. Writing–original draft: KIPY, GASP. Writing–review & editing: SSD, AJFR. All authors read and approved the final manuscript.

