## Supplemental Table 1 for "Association between intraoperative alactic base excess and postoperative acute kidney injury after liver transplantation: a retrospective cohort study"

| Variable | aRR (95%CI) | p value |
|---|---|---|
| Age | 0.99 (0.966 to 1.02) | 0.449 |
| Sex | 1.17 (0.655 to 2.09) | 0.594 |
| Body mass index | 1.01 (0.950 to 1.08) | 0.674 |
| MELD score | 1.04 (0.988 to 1.09) | 0.144 |
| Cold ischemia time | 1.00 (0.807 to 1.25) | 0.973 |
| Anhepatic phase duration | 1.02 (0.786 to 1.31) | 0.904 |
| Alactic base excess | 0.704 (0.419 to 1.18) | 0.185 |

Supplemental Table 1: Modified Poisson regression with robust variance. Alactic base excess as a dichotomous variable ($\leq -2.5$ mmol/L). MELD: Model for End-Stage Liver Disease; aRR: adjusted relative risk; 95% CI: 95% confidence interval.
