## Supplemental Table 2 for "Association between intraoperative alactic base excess and postoperative acute kidney injury after liver transplantation: a retrospective cohort study"

| Variable | aRR (95%CI) | p value |
|---|---|---|
| Age | 0.993 (0.970 to 1.02) | 0.565 |
| Sex | 1.07 (0.588 to 1.95) | 0.822 |
| Body mass index | 1.01 (0.944 to 1.08) | 0.756 |
| MELD score | 1.03 (0.984 to 1.09) | 0.185 |
| Cold ischemia time | 1.03 (0.843 to 1.27) | 0.749 |
| Anhepatic phase duration | 1.01 (0.772 to 1.32) | 0.954 |
| Alactic base excess | 0.904 (0.711 to 1.15) | 0.411 |
| ABExanhepatic_phase | 1.00 (0.913 to 1.10) | 0.979 |

Supplemental Table 2: Modified Poisson regression with robust variance, including an interaction between alactic base excess and anhepatic phase duration. MELD: Model for End-Stage Liver Disease; aRR: adjusted relative risk; 95% CI: 95% confidence interval.
