## Supplemental Table 3 for "Association between intraoperative alactic base excess and postoperative acute kidney injury after liver transplantation: a retrospective cohort study"

| Creatinine-based KDIGO stage | n (%) | ABE, mmol/L, median (IQR) |
|---|---|---|
| KDIGO 0 | 38 (54.3) | 0.75 (−1.52 to 2.85 |
| KDIGO 1 | 18 (25.7) | −1.20 (−2.85 to 0.18) |
| KDIGO 2 | 8 (11.4) | −3.30 (−3.32 to −0.90) |
| KDIGO 3 | 6 (8.6) | 1.65 (−0.50 to 2.38) |
| | Kruskal–Wallis p=0.028 | |

Supplemental Table 3: Distribution of alactic base excess according to creatinine-based KDIGO stage
