## Supplemental Table 4 for "Association between intraoperative alactic base excess and postoperative acute kidney injury after liver transplantation: a retrospective cohort study"

| Exposure | Model | RR/aRR (95%CI) | p-value |
|---|---|---|---|
| ABE, per 1 mmol/L increase | Unadjusted | 0.90 (0.73–1.10) | 0.30 |
| ABE, per 1 mmol/L increase | Adjusted | 0.98 (0.78–1.23) | 0.87 |
| ABE ≤−2.5 vs >−2.5 mmol/L | Adjusted | 1.51 (0.58–3.94) | 0.40 |

Supplemental Table 4: Post hoc association between alactic base excess and severe acute kidney injury. ABE: alactic base excess; RR: risk ratio; aRR: adjusted risk ratio; 95%CI: 95% confidence interval.
